# SARS-CoV2 serology assays: utility and limits of different antigen based tests through the evaluation and the comparison of four commercial tests

**DOI:** 10.1101/2021.11.19.21266615

**Authors:** Mariem Gdoura, Habib Halouani, Mehdi Mrad, Sahli Donia, Wafa Chamsa, Manel Mabrouk, Kamel Ben Salem, Nahed Hogga, Henda Triki

## Abstract

**Introduction:** SARS-CoV2 serology testing is multipurpose provided to choose an efficient test. We evaluated and compared 4 different commercial serology tests, three of them had the Food and Drug Administration (FDA) approval. Our goal was to provide new data to help to guide the interpretation and the choice of the serological tests.

**Methods:** Four commercial tests were evaluated: Cobas®Roche®(total anti-N antibodies), VIDAS®Biomerieux®(IgM and IgG anti-RBD antibodies), Mindray®(IgM and IgG anti-N and anti-RBD antibodies) and Access®Beckman Coulter®(IgG anti-RBD antibodies). Were tested: a positive panel (n=72 sera) obtained from COVID-19 confirmed patients and a negative panel (n=119) of pre-pandemic sera. Were determined the analytical performances and was drawn the ROC curve to assess the manufacturer’s threshold.

**Results:** A large range of variability between the tests was found. Mindray®IgG and Cobas® tests showed the best overall sensitivity 79,2%CI95%[67,9-87,8]. Cobas® showed the best sensitivity after D14; 85,4%CI95%[72,2-93,9]. The best specificity was noted for Cobas®, VIDAS®IgG and Access® IgG(100%CI95%[96,9-100]). Access® had the lower sensitivity even after D14 (55,5% CI95%[43,4-67,3]). VIDAS®IgM and Mindray®IgM tests showed the lowest specificity and sensitivity rates. Overall, only 43 out of 72 sera gave concordant results (59,7%). Retained cut-offs for a significantly better sensitivity and accuracy, without altering significantly the specificity, were: 0,87 for Vidas®IgM(*p*=0,01), 0,55 for Vidas®IgG(*p*=0,05) and 0,14 for Access®(*p*<10^−4^).

**Conclusion:** Although FDA approved, each laboratory should realize its own evaluation for commercial tests. Tests variability may raise some concerns that seroprevalence studies may vary significantly based on the used serology test.

## Introduction

The severe acute respiratory syndrome coronavirus 2 (SARS-CoV2) is an emerging virus that was first reported in December 2019 in Wuhan, China (1,2). Rapidly, the virus has spread across the globe and has become a major public health concern. In 11 March 2020, the World Health Organisation (WHO) announced the COVID-19 disease as a pandemic (3). To date, millions of infections by SARS-CoV-2 and hundreds of thousands of deaths have been attributed to the COVID-19. According to WHO COVID-19 dashboard as of 15 September 2021, the total of infections and deaths numbers are 225 680 357 and 4 644 740, respectively (4). Molecular testing by real time PCR is the angular stone in the diagnosis of the COVID-19 as it is playing a crucial role in testing, monitoring and contact tracing (5). However, as screening indications are mainly limited to symptomatic patients, documented cases represent probably only the visible part of the iceberg. For these reasons, other diagnostic methods are needed to better estimate COVID-19 spread (6,7). Serology testing suits ideally for this purpose as the detection of specific anti SARS-CoV2 antibodies offers valuable information about previous contact with the virus, helps to assess the herd immunity at a large or specific population, and recently, have decisive role to monitor vaccinated patients (8–10). A worldwide laboratories and companies competition was launched soon after the virus emergence, to develop efficient serology tests with good sensitivity and specificity, ease to use, rapid result and reasonable cost-effectiveness balance (11). Today many different tests are commercially available: enzyme-linked immune-sorbent assay (ELISA), enzyme-linked fluorescent assay (ELFA), eletrochimiluminescent assay (ECLIA). These tests detect different isotypes; IgM, IgG, IgA, total antibodies and use different antigens; the full spike glycoprotein or sub-units S1 and S2, the receptor-binding protein (RBD) or nucleoprotein N (11–17). International health authorities such as the Food and Drug Administration and the World Health Organization agreed to grant what they called an emergency use authorization (FDA-EUA) and an emergency use list (WHO-EUL), respectively (18–20). Nowadays, serology has become widely used with many indications(21,22). However, some tests may lack sufficient clinical evaluation which made specialists establishing their own evaluation and sharing concerns toward their performances(12,14–17).

In the present study we conducted a head-to-head comparison of 4 different commercial serology tests targeting either the N protein or the RBD protein or both of them; three of them had an FDA-EUA. Our goal was to provide experimental data which help to guide the choice of the serological tests according to their indication, as well as the interpretation of the serology profiles.

## Methods

The study was done in the Laboratory of Virology of the Institut Pasteur of Tunis, Tunisia and was approved by institutional review boards at the Institut Pasteur of Tunis. The selection of samples followed the guidelines of the French “*Centre National de Reference des Virus des Infections Respiratoires*” published on December 4^th^ 2020 (23), i.e. the evaluation needs at least 50 true positive sera and at least 50 true negative sera. A positive panel was composed of a total of 72 unique, non duplicated serum samples obtained from COVID-19 confirmed patients on the basis of a positive RT-PCR on nasopharyngeal swab. Samples were collected from the first day (D0) until 162 day (D162) after molecular confirmation. Serum samples included 29 sera collected from D1 to D14, 16 sera from D16 to D30, 14 sera from D31 to D60 and 13 sera after D60 (until D162). Along with the positive panel, a negative panel was included in the study; it is composed of 119 pre-pandemic sera collected before December 2019 and served as negative controls. Four commercial tests were evaluated: Cobas®Roche® (ECLIA) detecting total anti-N antibodies, VIDAS®Biomerieux® (ELFA) detecting specific IgM and IgG anti-RBD antibodies, Mindray® (CLIA) detecting specific IgM and IgG anti-N and anti-RBD antibodies and Access®Beckman Coulter® (CLIA) detecting specific IgG anti-RBD antibodies. All sera, the 72 from confirmed cases and the 119 pre-pandemic sera were tested by the 4 tests and manipulations were carried out according to the manufacturers’ instructions. The intrinsic characteristics and the manufacturer’s performances of each test are summarized in Table 1.

**Table 1:** Characteristics of the automated analyzers used for SARS-CoV-2 antibodies detection.

| Automated Analyzer | Certifications and EUA | detection Method | antibody detected | targeted antigen | sample volume | cut-off | result interpretation | reported sensitivity | reported specificity |
| --- | --- | --- | --- | --- | --- | --- | --- | --- | --- |
| **Vidas®Bio**<br>**mérieux®** | CE-IVD,<br>FDA-EUA | ELFA | IgM<br>IgG | S1 RBD | 100 uL<br>including the<br>dead volume | 1 for<br>both | <1 Negative<br>> or = 1 Positive | IgG: n=71 out of 120, PPA 59,2%<br>CI95%[49,8-68] 100%<br>[66.4; 100.0]<br>IgM: n=69 out of 111, PPA 62,2%<br>CI95%[52,5-71,2], > 8 days 100%<br>[63.1; 100.0] | IgG: 99.9%<br>[99.4-100.0] ,<br>IgM: 99.4%<br>[97.7-99.9]. |
| **CL-**<br>**900i®Mind**<br>**ray®** | CE-IVD | CLIA | IgM<br>IgG | S and N<br>protein | 10 uL with a<br>minimum of<br>100 uL of<br>dead volume | 1 for<br>IgM<br>1 for<br>IgG | <1 Negative<br>> or = 1 Positive for<br>IgM and <10<br>Negative<br>> or = 10 Positive | 82.22% | 87.60% |
| **Elecsys®R**<br>**oche®** | CE-IVD,<br>FDA-EUA,<br>WHO-EUL | ECLIA | Total<br>antibodies:<br>IgG+++,<br>IgM and<br>IgA | N<br>protein | 10 uL with a<br>minimum of<br>100 uL of<br>dead volume | 1 | <1 Negative<br>> or = 1 Positive | 100% [88,1-100] after 14 days | 99.8 %. [99,7-99,9] |
| **Access®**<br>**Beckman®** | CE-IVD,<br>FDA-EUA | CLIA | IgG | S1 RBD | 10 uL with a<br>minimum of<br>100 uL of<br>dead volume | 1 | <1 Negative<br>> or = 1 Positive | 100% [93,8-100] after 18 days | 99.8 %. [99,4-99,9] |

Were determined: the overall sensitivity, the sensitivity after D14, the specificity, the positive predictive value PPV, the negative predictive value NPV, the area under the curve (AUC) and the correlation between the results (in ratio index/cut-off index (ratios)) with the days after COVID-19 confirmation. Based on the obtained ratios, Receiving operating characteristic (ROC) curve for each test has been drawn using the obtained ratios, in order to assess the manufacturer’s threshold to achieve the best performances. Finally, the 4 tests were compared between each other and pooled results of different tests were statistically studied.

### Statistical Analysis

PPV and NPV were calculated using the FDA calculator available on its website, accessed 22 July 2021 and arbitrary fixed the prevalence of the disease at 5%. Using RT-PCR as the reference standard, sensitivity, specificity, AUC were calculated to assess the performance of each assay and T-test was used to compare AUC for each test. The optimal cutoff point was selected based on the point with the highest Youden index J. Agreement was determined between all the tests, two by two, using the Cohen’s Kappa statistic test. Correlation between ratios was calculated by determining the Pearson Coefficient r. The significance level was set at 5%, and a 95% confidence interval (CI95%) was reported for each measure. All calculations were performed using MEDCALC®V18.2.1.

## Results

The performances of each test were evaluated by calculating the sensitivity, specificity, PPV, NPV and AUC. All experimental results are shown in Table 2. For IgM tests, VIDAS®IgM and Mindray®IgM tests showed the lowest specificity and sensitivity rates for all the sera and those collected after D14. The PPV and NPV were also the lowest. False positive tests were obtained for 7 pre-pandemic patients for Vidas®IgM: 3 patients having rheumatoid factors and 4 patients positive for *Herpessimplex virus;* the ratios ranged from 1,08 to 13,94. False positive tests were obtained for 3 pre-pandemic patients for Mindray®IgM: 2 patients having auto-immune disease and 1 patient positive for *Herpessimplex virus;* the ratios ranged from 1,94 to 4,97. These two tests had good and similar accuracies (*p*=0,587). For IgG and total antibody tests, Mindray®IgG and Cobas® tests showed the best overall sensitivity 79,2% CI95%[67,9-87,8] (57 positive out of 72 true positive). Cobas® showed the best sensitivity after D14 85,4%CI95%[72,2-93,9]. For the other tests, the sensitivity increased considerably after D14 except for Access® (Table 2). The best specificity and PPV were noted for Cobas®, VIDAS®IgG and Access® IgG: 100%CI95%[96,9-100] (119 negative out of 119 true negative). Mindray®IgG was slightly less specific (95,8% CI95%CI95%[90,5-98,6]), and PPV was low 49,8%CI95%[29,4-70,2]. Positive results were obtained for 5 pre-pandemic patients: 1 having rheumatoid factors, 2 patients positive for *Herpessimplex virus* and 2 pregnant women, ratios ranged from 1,3 to 3,8. Considering accuracy, Cobas®, VIDAS®IgG and Mindray®IgG had very good and similar accuracy (paiwise comparison of ROC curves for the 3 combinations *p*>0,05). However, Access® had an accuracy of 0,778CI95%[0,712-0,835] which is good but statistically lower than the other tests (*p*=0,587). ROC curves for each test were drawn based on the obtained ratios (Figure 1) as shown in Tables 3. Retained cut-offs for a significantly better sensitivity and accuracy, without altering significantly the specificity, were: 0,87 instead of 1 for Vidas®IgM (*p*=0,01), 0,55 instead of 1 for Vidas®IgG (*p*=0,05) and 0,14 instead of 1 for Access® (*p*<10^−4^). For Cobas® and Mindray®IgM and IgG, the new proposed cut offs did not give better analytical performances than the original cut-offs (*p*>0,05, Table 3).

**Table 2:** Experimental analytical performances for the 4 automated analyzers according to the manufacture criteria and to the new proposed criteria.

|  | false - | false –<br>≥ 14 days | true + | true +<br>≥ 14 days | Sensitivity %<br>CI95% | sensitivity ≥<br>14 days %<br>CI95% | false + | true - | Specificity %<br>CI95% | AUC CI95% | VPP<br>% CI95% | VPN<br>% CI95% |
| --- | --- | --- | --- | --- | --- | --- | --- | --- | --- | --- | --- | --- |
| **Vidas® IgM** | 35 | 29 | 37 | 19 | 51,4<br>[39,3-63,3] | 39,6<br>[25,7-54,7] | 7 | 112 | 94,1<br>[88,2-97,6] | 0,728<br>[0,659-0,789] | 31,5<br>[17,8-49,4] | 97,4<br>[96,6-97,9] |
| **Vidas® IgG** | 17 | 8 | 55 | 40 | 76,4<br>[64,9-85,6] | 83,3<br>[69,8-92,5] | 0 | 119 | 100<br>[96,9-100] | 0,882<br>[0,828-0,924] | 100 | 98,7<br>[98,1-99,2] |
| **Cobas®** | 15 | 7 | 57 | 41 | 79,2<br>[68-87,8] | 85,4<br>[72,2-93,9] | 0 | 119 | 100<br>[96,9-100] | 0,896<br>[0,844-0,935] | 100 | 98,9<br>[98,3-99,3] |
| **Access®** | 32 | 20 | 40 | 28 | 55,5<br>[43,4-67,3] | 41,7<br>[27,6-56,8] | 0 | 119 | 100<br>[96,9-100] | 0,778<br>[0,712-0,835] | 100 | 97,7<br>[97-98,2] |
| **Mindray® IgM** | 40 | 32 | 32 | 16 | 44,4<br>[32,7-56,6] | 66,7<br>[51,6-79,6] | 3 | 116 | 97,5<br>[92,8-99,5] | 0,710<br>[0,640-0,773] | 48,1<br>[22,7-74,5] | 97<br>[96,4-97,6] |
| **Mindray® IgG** | 15 | 8 | 57 | 40 | 79,2<br>[68-87,8] | 83,3<br>[69,7-92,5] | 5 | 114 | 95,8<br>[90,5-98,6] | 0,875<br>[0,819-0,918] | 49,8<br>[29,4-70,2] | 98,8<br>[98,2-99,3] |

**Figure 1:**
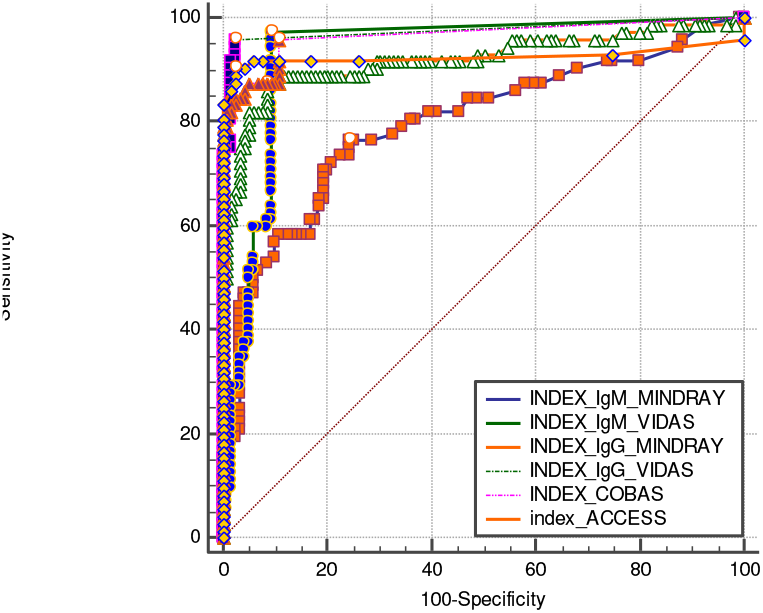
ROC curves for the 4 tests.

**Table 3:** Results of the ROC analysis for new cut-offs.

| | Original cut-off | Proposed cut-off | sensitivity%<br>CI95% | sensitivity ≥ 14<br>days% CI95% | specificity%<br>CI95% | AUC CI95% | *P* |
| --- | --- | --- | --- | --- | --- | --- | --- |
| **Vidas® IgM** | 1 | >0,87 | 59,7<br>[47,5-71,1] | 52<br>[37,2 66,7] | 94,1<br>[88,3-97,6] | 0,767<br>[0,703-0,827] | 0,01 |
| **Vidas® IgG** | 1 | >0,55 | 84,7<br>[74,3-92,1] | 91,2<br>[80 97,7] | 98,3<br>[94,1-99,8] | 0,922<br>[0,875-0,956] | 0,05 |
| **Cobas®** | 1 | >0,725 | 81,94<br>[71,1-90] | 85,4<br>[72,2 93,9] | 99,2<br>[95,4-100] | 0,906<br>[0,855-0,943] | 0,36 |
| **Access®** | 1 | >0,14 | 83,3<br>[72,7-91,1] | 83,3<br>[69,8 92,5] | 100<br>[96,9-100] | 0,917<br>[0,868-0,952] | $<10^{-4}$ |
| **Mindray® IgM** | 1 | >0,83 | 47,2<br>[35,3-59,3] | 37,5<br>[23,9 52,6] | 95,8<br>[90,5-98,6] | 0,715<br>[0,645-0,778] | 0,63 |
| **Mindray® IgG** | 10 | >7,94 | 81,9<br>[72,7-91,9] | 87,5<br>[74,7 95,2] | 94,1<br>[88,3-97,6] | 0,887<br>[0,834-0,928] | 0,34 |

All experimental results obtained for the positive panel were summarized in Table 4 including concordance and discordance between the tests and in connexion also with the days after confirmation. Overall, 43 ou of 72 sera gave concordant results (59,7% concordance). Of them, 35 were positive, sampled between D4 and D140 and 8 were negative, sampled between D0 and D60. Discordant results represented 40,3% of the panel (29 out of 72). They were divided into 3 groups: the first group (n=17) contains samples that were positive by 3 tests over 4, Access® failed to detect 13 samples collected between D6 and D90 and Cobas® failed to detect 4 samples collected between D8 and D39 d. The second group (n=11) included samples that were positive by only 2 tests over 4. We obtained positive results by Mindray® in concordance with another test (Vidas® or Cobas®) in 7 cases out of 11. The third group contains one sample collected at D7 detected positive by only Vidas® and negative by the other tests.

**Table 4:** Concordance and discordance between the 4 evaluated tests results for the positive panel (n=72)

| Group | N | VIDAS® IgG<br>and/or IgM | Cobas® | Access ® | Mindray ® IgG<br>and/or IgM | DAYS* |
| --- | --- | --- | --- | --- | --- | --- |
| C1 | 35 | + | + | + | + | 25 [4-140] |
| C2 | 8 | - | - | - | - | 14 [0-60] |
| 3 positive<br>tests over 4<br>n=17 | 0 | + | + | + | - | NA |
|  | 13 | + | + | - | + | 21 [6-90] |
|  | 4 | + | - | + | + | 20 [8-39] |
|  | 0 | - | + | + | + | NA |
| 2 positive<br>tests over 4<br>n=11 | 3 | + | + | - | - | 39, 90, 162 |
|  | 1 | + | - | + | - | 17 |
|  | 1 | + | - | - | + | 9 |
|  | 0 | - | + | + | - | NA |
|  | 6 | - | + | - | + | 23 [15-86] |
|  | 0 | - | - | + | + | NA |
| 1 positive<br>test over 4<br>n=01 | 1 | + | - | - | - | 7 |
|  | 0 | - | + | - | - | NA |
|  | 0 | - | - | + | - | NA |
|  | 0 | - | - | - | + | NA |

To further understand the differences between the tests results, we determined the agreement between the results and the correlation between the ratios. In this section, the comparison was focused on the isotype and on the antigen. For the IgM tests, Vidas® and Mindray® present moderate agreement (k=0,570) and weak correlation (r=0,484). For the other tests, the results of the agreement and correlation are grouped in Figure 2. For tests detecting exclusively antibodies against RBD, Vidas®IgG and Access®, concordance was important and correlation is positive and strong. For Cobas®, the test that detects antibodies against N exclusively, agreement was perfect with Vidas® and important with Access® and correlation is positive and moderately strong with Vidas® but negative with Access®. Considering the Mindray®, the test that detects both N and RBD specific antibodies, it presents a perfect agreement with all tests except Access® and a positive correlation with all tests.

**Figure 2:**
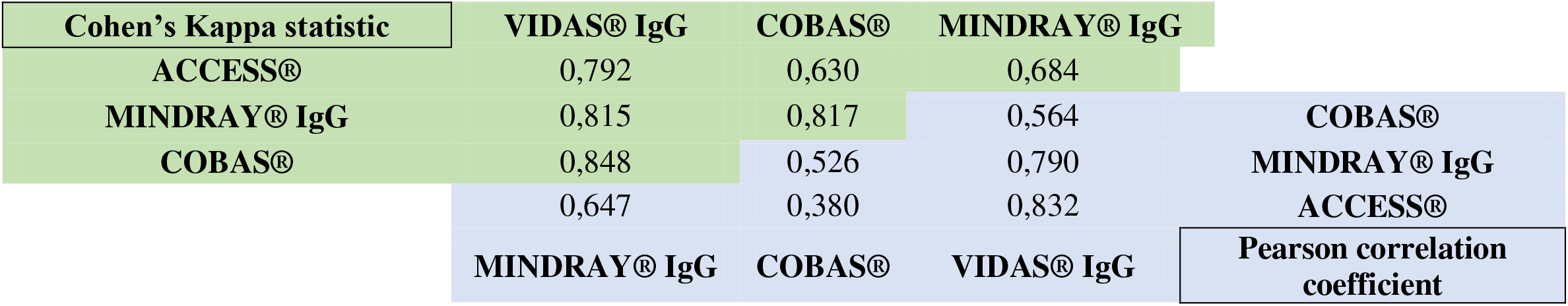
Agreement between qualitative results and correlation between COI/COI cut-offs ratios.

Figure 3 shows the scatterplots of the 4 tests’ ratios against d. Figure 3.a shows the distribution of the IgM tests indexes (Vidas® and Mindray®), which is heterogenous et does not fit a specific pattern however it is obvious that high indexes were obtained during the first 21 days after infection. Figure 3.b illustrates the distribution of total antibodies and IgG antibodies tests indexes (Cobas®, Vidas®, Mindray® and Access®) and shows that there is no correlation with days after infection. However, for Cobas®, ratios seem to be increasing over days. Different test results combinations’ were studied by pooling the obtained results. No improvement of performances was noted for [Cobas®, *versus* Cobas® and Vidas® IgG and IgM], for [Cobas® *versus* Mindray®IgM+IgG], for [Vidas®IgG+IgM *versus* Mindray®IgM+IgG] (pairwise comparison of ROC curves *p*>0,05). Only the combination Vidas®IgG+IgM and Cobas® was consistenly more accurate than Mindray®IgM and IgG (pairwise comparison of ROC curves *p*=0,0399).

**Figure 3.**
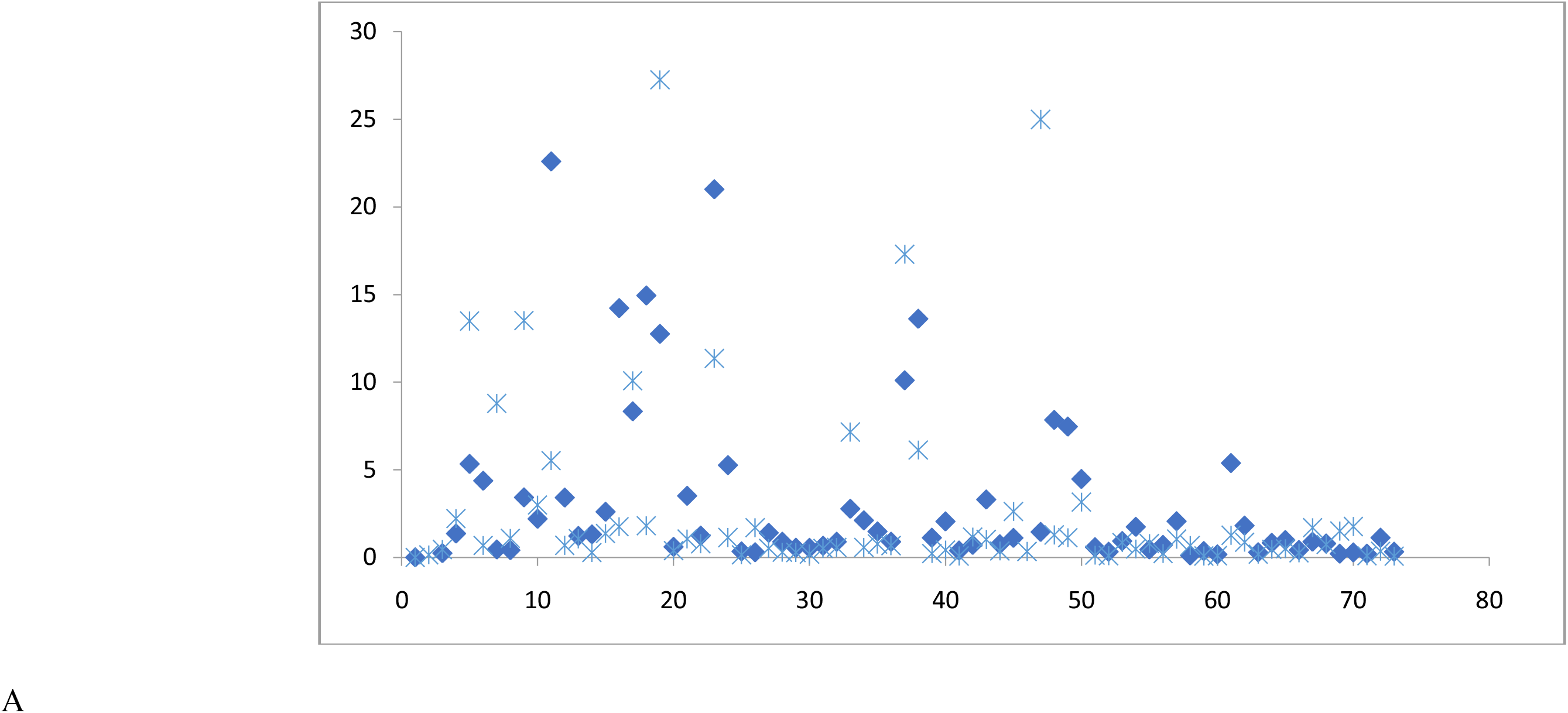
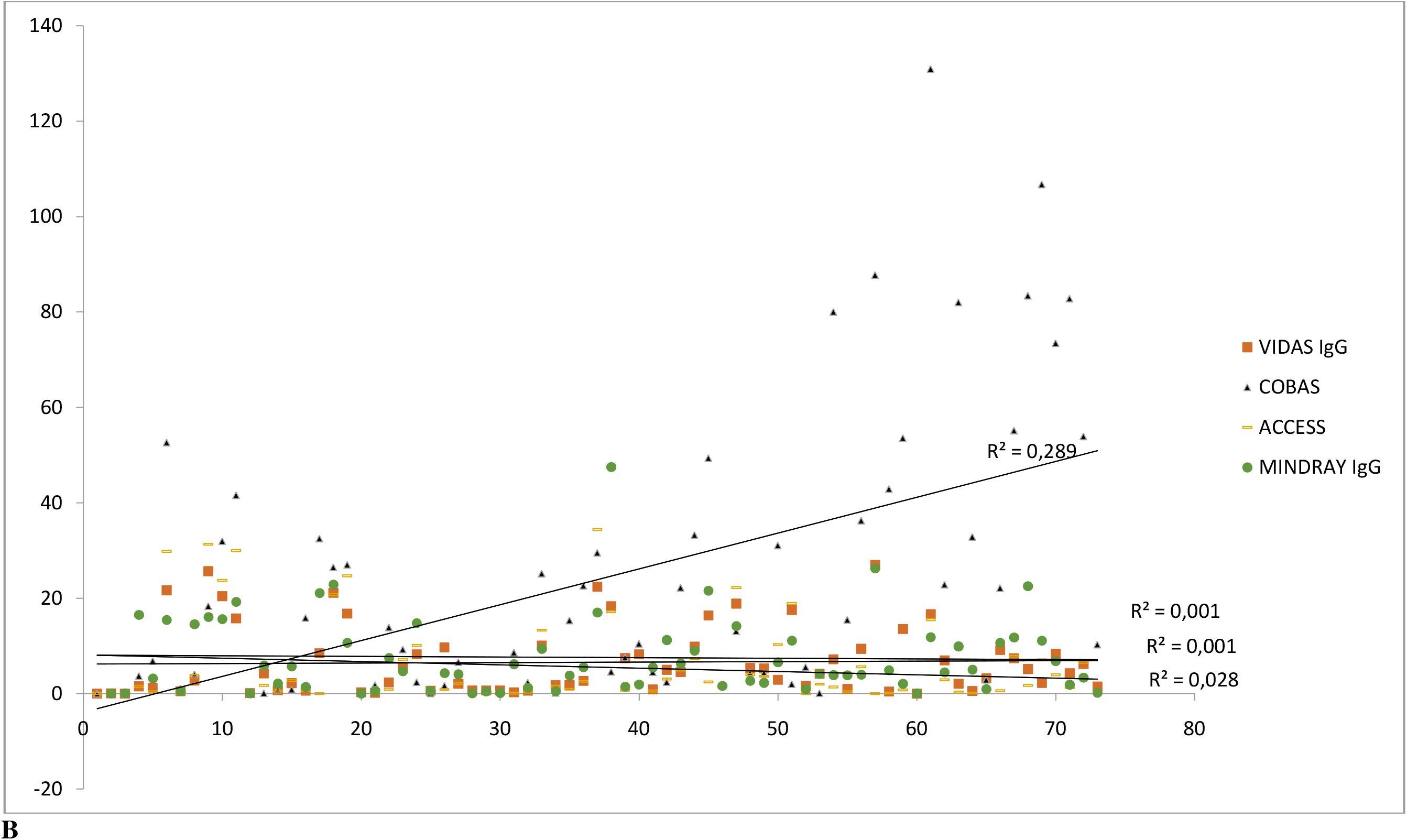
Scatterplot tests indexes plotted against day of RT-PCR positive results. A: IgM tests, B: other tests.

## Discussion

SARS-CoV2 serology tests were developed and optimized in a record time after the virus emergence. Thanks to the the softened authorization procedure, they were rapidly commercialized and used worldwide(18–20). In this study, we evaluated and compared 4 serology commercial automated tests: Cobas®Roche® (ECLIA) detecting total anti-N antibodies, IgG+++ (anti-N), VIDAS®Biomerieux® (ELFA) detecting specific IgM and IgG anti-RBD antibodies, Mindray® (CLIA) detecting specific IgM and IgG anti-N and anti-RBD antibodies and Access®Beckman Coulter® (CLIA) detecting specific IgG anti-RBD antibodies. Our evaluation revealed a gap between claimed and experimental analytical performances in terms of sensitivity and specificity and, accordingly, we propose new analytical criteria. In addition, the comparison between the evaluated tests showed a significant divergence between the obtained qualitative results in 40,3% of the positive tested sera (29 out of 72). Our findings suggest that the most sensitive test, after D14 is Cobas® (85,4%IC95%[72,2-93,9]) which detects high ratios until 4 months after primo-infection. Besides, we found that combining RBD and N tests from different tests gives the best accuracy.

We tested 72 RT-PCR confirmed patients and 119 pre-pandemic sera. Our work stands out from the rest of the literature by studying tests of different antigen and having international approved certificate, by proposing new significant cut-offs to improve the analytical performances and by a deep assessment of the origins behind discordances of the obtained results as well as the discussion of the utility and the limits of each test. Thus, our work provides original and helpful data serving the health care professionals in their routine practice. Even our panel is not too large, the number of tests is not big and the impact of the disease severity was not studied, our results are extrapolable given that the panel is representative (from D0 until D162), the tests are diversified (different antigens and isotypes) and the conclusions are applicable for a diagnosis laboratory receiving all type of indications. The evaluation of commercial tests was widely reported for SARS-CoV2 virus as well as for other pathogens. International recommendations were published by several scientific socities and instances such as *Haute Autorité Sanitaire France* HAS, FINDXX and PHE and Health Canada in order to harmonize the criteria of validation of the tests (23–26). In our study, the evaluation of all tests gave lower performances than the claimed ones and did not respond to the HAS validation criteria, the most flexible one, in terms of sensitivity (Table 1,2). According to the HAS, the sensitivity of detecting IgG and total antibodies must exceed 90% after D14 from disease onset while for IgM antibodies, the sensitivity must exceed 90% after D7. All tests were studied specifically after D14, based on a large review published by Cochrane on 15976 samples which found that all the results for IgG, IgM, IgA, total antibodies showed low sensitivity during the first week after the symptoms onset (all less than 30.1%), it rises in the second week and reaches its highest values in the third week (36).

In our series, the best sensitivity after D14 were the one of Cobas® (85,4%CI95%[72,2-93,9]) followed by Vidas®IgG and Mindray®IgG (83,3%CI95%[69,8-92,5]) and Access® came in the last position(55,5% CI95% [43,4-67,3]) (Table 2). For IgM detection Mindray® and Vidas® had very low sensitivity even after D14. High sensitivity for Cobas® confirms the findings of other authors (12,15,16,27). This could be explained, first, by the used antigen which is exclusively the N protein, known to be the most abundantly expressed immune-dominant protein (21). Second is the ability of Cobas® to detect all immunoglobulin classes and such data was reported for Siemens Atellica® that detects total antibodies anti-glycoprotein S1(28). Third is the ECLIA Elecsys® technology developed by Roche® which is highly efficient regardless of the measured analyte(29,30). For the other tests, such unsatisfactory sensitivities were reported by other studies for the same tests. Similar low sensitivities for the Vidas® test were reported by Younes et al. (88,3 for Vidas®IgG after D21) and by Wolf et al. (over all sensitivity of 64,9%CI95%[55,2-73,7] for Vidas®IgM and 73%CI95%[63,7-81] for Vidas®IgG) (31,32). Padoan et al. also reported a sensitivity of 86.4 (77.0-93.0) for both Mindray®IgM and IgG tests but a new Mindray® generation would give much better performances: 99% and 96% from D1 to D41 for IgG and IgM, respectively (33,34). This new version of Mindray® was not available at the study writing time and merits to be evaluated. Access® showed very low sensitivity for IgG detection (55,5%CI95%[43,4-67,3]) and this sensitivity did not increase after D14. Other authors reported similar results for Access® 39.6%CI 95%[32.5–47.3%] and 69% CI95%[59.0-77.9] (15,35). Beckman® has developed a new Access® test allows semi quantified detection of antibodies against RBD and that has obtained the FDA-EUA and thus merits to be evaluated.

Regarding the specificity, Vidas®IgM and Mindray®IgM and IgG tests gave positive signals for few pre-pandemic sera and this was also reported by other authors (31,33,34). Cross reactivity with pre-pandemic auto-immune disease patients sera was previously reported(37).

In contrast, cross reactivity with pre-pandemic pregnant women sera, and patients positive for H*erpessimplex virus* is reported for the first time. Results of Vidas®IgM and Mindray®IgM and IgG should be interpreted with caution; indeed, PPV of these tests are less than 50%, which mean that half of tested patients are susceptible to be false positive. Here, PPV and NPV were calculated by the FDA calculator fixing the prevalence at 5%. Each coutry is invited to evaluate regularily the PPV according to the prevalence evolution. The tests Cobas®, Access® and Vidas® IgG are the extremely specific (100%IC95%[96,9-100]) and the PPV is 100%, as reported by other studies (16,27,31).

The pre-defined tests’ thresholds were experimentally optimized and adjusted for an improved sensitivity with very little loss in specificity. This approach is being widely used and reported by many authors for better interpretation of commercial tests, for COVID-19 tests as well as other pathogens (31,38,39). We found out that decreasing the cut-off signals for Vidas®IgM, Vidas®IgG and Access® improves significantly the sensitivity and the accuracy (Table3). As none of the tests propose a grey zone for borderline results, which is unusual, we propose that any results between the proposed cut-off and the original cut-off (i.e: [0,87 to 1] for Vidas®IgM, [0,55–1] for Vidas®IgG and [0,14–1] for Access®) should be retested or, better, the patient should be re-sampled after 10 to 15 days to follow the antibody kinetic. More generally, we suggest that any weak signal less than 2 times the cut-off, should be interpreted with caution.

Comparison between the four tests showed concordant results in 59,7% of the samples collected in confirmed cases (43 out of 72) among which, 8 were negative by all the 4 tests; they were sampled between D0 and D60, median=14. As the 4 different tests using different antigens, gave negative results, it is suggested that this negativity is inherent to the individuals. Indeed, this may be explained by either a late sero-conversion, or a rapid sero-reversion (40). Some authors has suggested that 5 to 10% of infected persons do not develop antibodies at all (41). A non negligible proportion of discordant results was found in 40,3% sera (n=29 out of 72). Access® was the test that fails the most to detect positive results (13 cases out of 29 discordant results, Table 4). For the rest of the tests, although general agreement between qualitative results is important (figure 2), they gave various discordant patterns.

A dominated discordant pattern is interesting, it is about positive results for Mindray® (24 cases out of 29 cases), indicating that a combination of N and RBD antigens would increase the number of true positives sera. Indeed, Mindray® is not an FDA-EUA test, but was introduced in this study for its originality as it is multiplex (N+RBD). We demonstrated that Mindray®IgM and IgG offers similar good sensitivity to Cobas®, but the combinaison [Cobas®+Vidas®IgM+IgG] exceeds Mindray® IgM+IgG in accuracy. So, a two steps strategy starting by testing Cobas® then Vidas®IgM+IgG could improve significantly the sensitivity, and offers separate comprehension of antibodies specificity.

Questions regarding the magnitude and the longevity of the antibody response remain unanswered. Many literature reviews tried to propose a general kinetic of antibodies and recognize a big variability between individuals and proportionality with COVID-19 severity (42,43). In our study, Figures 3.a showed that scatter plots of the two IgM tests are high and condensed among the first 3 weeks, suggesting that their detection is in line with an ongoing or acute infection. However, we found that IgM may still detectable even until D162, Regarding IgG (Figure 2 and 3.b) anti RBD antibodies follow the same decay contrasting with the anti-N that persists positive with high ratios for longer time. This is explained by half life time for IgG anti RBD which is 49 days versus the half time of the IgG anti-N is 75 days (43).

In conclusion, although serological assays do not replace molecular tests in diagnosing active infection, they are multipurpose provided to choose the most efficient test and to properly interpret the results. We characterized the performance of four commercial antibody platforms and found out and explained the variability between them. Although FDA approved, each laboratory should realize its own evaluation for commercial tests, and health professionals should be aware about false negative rate before 14 to 21 days after primo-infection. Finally, this variability may raise some concerns that seroprevalence studies may vary significantly based on the used serology test.

## Data Availability

All data produced in the present study are available upon reasonable request to the authors

## Acknowledgment

We would like to thank Dr Malika Ben Ahmed from the laboratory of immunology of the Institut Pasteur of Tunis, Tunisia, for providing pre-pandemic sera positive for rheumatoid factors.

## References

1. Zhou P, Yang X-L, Wang X-G, Hu B, Zhang L, Zhang W, et al. A pneumonia outbreak associated with a new coronavirus of probable bat origin. Nature. mars 2020;579(7798):270–3.

2. The species Severe acute respiratory syndrome-related coronavirus: classifying 2019-nCoV and naming it SARS-CoV-2. Nat Microbiol. 2020;5(4):536–44.

3. Chronologie de l’action de l’OMS face à la COVID-19 [Internet]. [cited 25 sept 2021]. available on : https://www.who.int/fr/news/item/29-06-2020-covidtimeline

4. WHO Coronavirus (COVID-19) Dashboard [Internet]. [cited 25 sept 2021]. Available on: https://covid19.who.int

5. Esbin MN, Whitney ON, Chong S, Maurer A, Darzacq X, Tjian R. Overcoming the bottleneck to widespread testing: a rapid review of nucleic acid testing approaches for COVID-19 detection. RNA N Y N. juill 2020;26(7):771–83.

6. Nishiura H, Kobayashi T, Miyama T, Suzuki A, Jung S, Hayashi K, et al. Estimation of the asymptomatic ratio of novel coronavirus infections (COVID-19). Int J Infect Dis. mai 2020;94:154–5.

7. Van Caeseele P, Bailey D, Forgie SE, Dingle TC, Krajden M. SARS-CoV-2 (COVID-19) serology: implications for clinical practice, laboratory medicine and public health. CMAJ Can Med Assoc J. 24 août 2020;192(34):E973–9.

8. Sidiq Z, Hanif M, Dwivedi KK, Chopra KK. Benefits and limitations of serological assays in COVID-19 infection. Indian J Tuberc. déc 2020;67(4):S163–6.

9. Dimeglio C, Herin F, Martin-Blondel G, Miedougé M, Izopet J. Antibody titers and protection against a SARS-CoV-2 infection. J Infect [Internet]. 20 sept 2021 [cited 25 sept 2021];0(0). Available on: https://www.journalofinfection.com/article/S0163-4453(21)00483-7/fulltext

10. Crotty S. Hybrid immunity. Science. 25 juin 2021;372(6549):1392–3.

11. Petherick A. Developing antibody tests for SARS-CoV-2. Lancet Lond Engl. 2020;395(10230):1101–2.

12. Schnurra C, Reiners N, Biemann R, Kaiser T, Trawinski H, Jassoy C. Comparison of the diagnostic sensitivity of SARS-CoV-2 nucleoprotein and glycoprotein-based antibody tests. J Clin Virol. août 2020;129:104544.

13. Younes N, Al-Sadeq DW, AL-Jighefee H, Younes S, Al-Jamal O, Daas HI, et al. Challenges in Laboratory Diagnosis of the Novel Coronavirus SARS-CoV-2. Viruses. juin 2020;12(6):582.

14. Nandakumar V, Profaizer T, Lozier BK, Elgort MG, Larragoite ET, Williams ESCP, et al. Evaluation of a Surrogate ELISA-Based Severe Acute Respiratory Syndrome Coronavirus 2 (SARS-CoV-2) cPass Neutralization Antibody Detection Assay and Correlation with IgG Commercial Serology Assays. Arch Pathol Lab Med. 28 juin 2021;

15. Tan SS, Saw S, Chew KL, Huak CY, Khoo C, Pajarillaga A, et al. Head-to-head evaluation on diagnostic accuracies of six SARS-CoV-2 serological assays. Pathology (Phila). éc 2020;52(7):770–7.

16. Riester E, Majchrzak M, Mühlbacher A, Tinguely C, Findeisen P, Hegel JK, et al. Multicentre Performance Evaluation of the Elecsys Anti-SARS-CoV-2 Immunoassay as an Aid in Determining Previous Exposure to SARS-CoV-2. Infect Dis Ther. 9 août 2021;1–17.

17. Murray MJ, McIntosh M, Atkinson C, Mahungu T, Wright E, Chatterton W, et al. Validation of a commercially available indirect assay for SARS-CoV-2 neutralising antibodies using a pseudotyped virus assay. J Infect. mai 2021;82(5):170–7.

18. Ravi N, Cortade DL, Ng E, Wang SX. Diagnostics for SARS-CoV-2 detection: A comprehensive review of the FDA-EUA COVID-19 testing landscape. Biosens Bioelectron. 1 oct 2020;165:112454.

19. Diagnostics laboratory emergency use listing [Internet]. [cited 25 sept 2021]. Available on: https://www.who.int/teams/health-product-and-policy-standards/about/regulation-and-prequalification

20. Health C for D and R. Emergency Use Authorizations for Medical Devices [Internet]. FDA. FDA; 2021 [cited 25 sept 2021]. Available on: https://www.fda.gov/medical-devices/emergency-situations-medical-devices/emergency-use-authorizations-medical-devices

21. CDC. Labs [Internet]. Centers for Disease Control and Prevention. 2020 [cited 25 sept 2021]. Available on: https://www.cdc.gov/coronavirus/2019-ncov/lab/resources/antibody-tests-guidelines.html

22. on behalf of the Canadian Public Health Laboratory Network (CPHLN) Serology Working Group, Charlton C, Kanji J, Tran V, Kus J, Gubbay J, et al. Practical guidance for clinical laboratories for SARS-CoV-2 serology testing. Can Commun Dis Rep. 7 mai 2021;47(04):171–83.

23. Arrêté du 3 décembre 2020 modifiant l’arrêté du 10 juillet 2020 prescrivant les mesures d’organisation et de fonctionnement du système de santé nécessaires pour faire face à l’épidémie de covid-19 dans le cadre de l’état d’urgence sanitaire - Légifrance [Internet]. [cited 25 sept 2021]. Available on: https://www.legifrance.gouv.fr/jorf/id/JORFTEXT000042607794

24. Canada H. COVID-19 serological testing devices: Notice on sensitivity and specificity values [Internet]. 2021 [cited 25 sept 2021]. Available on: https://www.canada.ca/en/health-canada/services/drugs-health-products/covid19-industry/medical-devices/testing/serological/notice-sensitivity-specificity-values.html

25. 20200427-COVID-19-IA-Evaluation-Synopsis.pdf [Internet]. [cited 25 sept 2021]. Available on: https://www.finddx.org/wp-content/uploads/2020/04/20200427-COVID-19-IA-Evaluation-Synopsis.pdf

26. Target Product Profile: antibody tests to help determine if people have recent infection to SARS-CoV-2: Version 2 [Internet]. http://GOV.UK. [cited 25 sept 2021]. Available on: https://www.gov.uk/government/publications/how-tests-and-testing-kits-for-coronavirus-covid-19-work/target-product-profile-antibody-tests-to-help-determine-if-people-have-recent-infection-to-sars-cov-2-version-2

27. Tan SS, Saw S, Chew KL, Wang C, Pajarillaga A, Khoo C, et al. Comparative Clinical Evaluation of the Roche Elecsys and Abbott Severe Acute Respiratory Syndrome Coronavirus 2 (SARS-CoV-2) Serology Assays for Coronavirus Disease 2019 (COVID-19). Arch Pathol Lab Med. 1 janv 2021;145(1):32–8.

28. Pflüger LS, Bannasch JH, Brehm TT, Pfefferle S, Hoffmann A, Nörz D, et al. Clinical evaluation of five different automated SARS-CoV-2 serology assays in a cohort of hospitalized COVID-19 patients. J Clin Virol. sept 2020;130:104549.

29. Douet T, Armengol C, Charpentier E, Chauvin P, Cassaing S, Iriart X, et al. Performance of seven commercial automated assays for the detection of low levels of anti-Toxoplasma IgG in French immunocompromised patients. Parasite Paris Fr. 2019;26:51.

30. Schmidt M, Jimenez A, Mühlbacher A, Oota S, Blanco L, Sakuldamrongpanich T, et al. Head-to-head comparison between two screening systems for HBsAG, anti-HBc, anti-HCV and HIV combination immunoassays in an international, multicentre evaluation study. Vox Sang. août 2015;109(2):114–21.

31. Younes S, Al-Jighefee H, Shurrab F, Al-Sadeq DW, Younes N, Dargham SR, et al. Diagnostic Efficiency of Three Fully Automated Serology Assays and Their Correlation with a Novel Surrogate Virus Neutralization Test in Symptomatic and Asymptomatic SARS-COV-2 Individuals. Microorganisms. 25 janv 2021;9(2):245.

32. Wolff F, Dahma H, Duterme C, Van den Wijngaert S, Vandenberg O, Cotton F, et al. Monitoring antibody response following SARS-CoV-2 infection: diagnostic efficiency of 4 automated immunoassays. Diagn Microbiol Infect Dis. 1 nov 2020;98(3):115140.

33. Padoan A, Cosma C, Zaupa P, Plebani M. Analytical and diagnostic performances of a high-throughput immunoassay for SARS-CoV-2 IgM and IgG [Internet]. 2020 nov [cited 25 sept 2021] p. 2020.11.20.20235267. Available on: https://www.medrxiv.org/content/10.1101/2020.11.20.20235267v1

34. Pieri M, Nuccetelli M, Nicolai E, Sarubbi S, Grelli S, Bernardini S. Clinical validation of a second generation anti-SARS-CoV-2 IgG and IgM automated chemiluminescent immunoassay. J Med Virol. avr 2021;93(4):2523–8.

35. COVID-19: laboratory evaluations of serological assays [Internet]. http://GOV.UK. [cited 25 sept 2021]. Available on: https://www.gov.uk/government/publications/covid-19-laboratory-evaluations-of-serological-assays

36. Deeks JJ, Dinnes J, Takwoingi Y, Davenport C, Spijker R, Taylor-Phillips S, et al. Antibody tests for identification of current and past infection with SARS-CoV-2. Cochrane Database Syst Rev [Internet]. 2020 [cited 25 sept 2021];(6). Available on: https://www.cochranelibrary.com/cdsr/doi/10.1002/14651858.CD013652/full

37. Jääskeläinen AJ, Kuivanen S, Kekäläinen E, Ahava MJ, Loginov R, Kallio-Kokko H, et al. Performance of six SARS-CoV-2 immunoassays in comparison with microneutralisation. J Clin Virol Off Publ Pan Am Soc Clin Virol. août 2020;129:104512.

38. Meyer B, Torriani G, Yerly S, Mazza L, Calame A, Arm-Vernez I, et al. Validation of a commercially available SARS-CoV-2 serological immunoassay. Clin Microbiol Infect. oct 2020;26(10):1386–94.

39. Performance characteristics of five immunoassays for SARS-CoV-2: a head-to-head benchmark comparison. Lancet Infect Dis. déc 2020;20(12):1390–400.

40. Milani GP, Dioni L, Favero C, Cantone L, Macchi C, Delbue S, et al. Serological follow-up of SARS-CoV-2 asymptomatic subjects. Sci Rep. 18 nov 2020;10(1):20048.

41. Petersen LR, Sami S, Vuong N, Pathela P, Weiss D, Morgenthau BM, et al. Lack of antibodies to SARS-CoV-2 in a large cohort of previously infected persons. Clin Infect Dis Off Publ Infect Dis Soc Am. 4 nov 2020;ciaa1685.

42. Huang AT, Garcia-Carreras B, Hitchings MDT, Yang B, Katzelnick LC, Rattigan SM, et al. A systematic review of antibody mediated immunity to coronaviruses: kinetics, correlates of protection, and association with severity. Nat Commun. 17 sept 2020;11(1):4704.

43. Piccoli L, Park Y-J, Tortorici MA, Czudnochowski N, Walls AC, Beltramello M, et al. Mapping Neutralizing and Immunodominant Sites on the SARS-CoV-2 Spike Receptor-Binding Domain by Structure-Guided High-Resolution Serology. Cell. 12 nov 2020;183(4):1024–1042.e21.

44. Zhang W, Du R-H, Li B, Zheng X-S, Yang X-L, Hu B, et al. Molecular and serological investigation of 2019-nCoV infected patients: implication of multiple shedding routes. Emerg Microbes Infect. 1 janv 2020;9(1):386–9.

45. WHO-2019-nCoV-lab-testing-2021.1-eng.pdf [Internet]. [cited 25 sept 2021]. Available on: https://apps.who.int/iris/bitstream/handle/10665/342002/WHO-2019-nCoV-lab-testing-2021.1-eng.pdf?sequence=1&isAllowed=y

